# Cohort Profile: National Institute for Health Research Health Informatics Collaborative: Hepatitis B Virus (NIHR HIC HBV) Research Dataset

**DOI:** 10.1101/2021.10.21.21265205

**Authors:** Tingyan Wang, David A Smith, Cori Campbell, Oliver Freeman, Zuzana Moysova, Theresa Noble, Kinga A Várnai, Steve Harris, Hizni Salih, Gail Roadknight, Stephanie Little, Ben Glampson, Luca Mercuri, Dimitri Papadimitriou, Christopher R Jones, Vince Taylor, Afzal Chaudhry, Hang Phan, Florina Borca, Josune Olza, Frazer Warricker, Luis Romão, David Ramlakhan, Louise English, Paul Klenerman, Monique Andersson, Jane Collier, Eleni Nastouli, Salim I Khakoo, William Gelson, Graham S Cooke, Kerrie Woods, Jim Davies, Eleanor Barnes, Philippa C Matthews

## Abstract

**Purpose:** The National Institute for Health Research (NIHR) Health Informatics Collaborative (HIC) was established to enable re-use of routinely collected clinical data across National Health Service (NHS) Trusts in the United Kingdom to support translational research. Viral hepatitis is one of the first five exemplar themes and hepatitis B virus (HBV) is the current focus of the theme. The NIHR HIC HBV dataset, derived from the central data repository of NIHR HIC viral hepatitis theme, aims to describe and characterise HBV infection in secondary care in the United Kingdom, and provides a resource for translational research.

**Participants:** The dataset comprises >5000 individuals (99% adults aged ≥18, 1% children aged <18) with chronic HBV (CHB) infection from five NHS Trusts across England, representing clinical data collected between August 1994 and August 2021.

**Findings to date:** Data on demographics, laboratory tests, antiviral treatment, elastography scores, imaging/biopsy reports, death information, and potential risk factors for liver disease have been collected. Data are captured by electronic patient record (EPR) systems, and records are updated prospectively as new results are added. This cohort profile describes the dataset in its current form. Among the adults, 55% are male, and the median age at index date (defined as the first recorded positive hepatitis B virus surface antigen (HBsAg) or HBV DNA in EPR systems) was 40 years (interquartile range [IQR]: 32-50). For those individuals with ethnicity reported, 30% were Asian, 24% were Black, 30% were White, and the remaining 16% were mixed or other ethnic groups. Currently, the median follow-up duration of the adult patients in this dataset was 5.0 (IQR: 2.7-7.5) years, with 9.3 (95% CI: 8.2-10.5) deaths per 1,000 person-years. We have already conducted several analyses using subsets of this dataset including an evaluation of distribution and trajectories of HBsAg and HBV viral load in CHB, reviewing the use of antiviral treatment, quantifying the burden of liver disease in the untreated population, and studying the use of laboratory biomarkers to improve stratification and surveillance.

**Future plans:** Longitudinal data collection is continuing, with the sample growing in size, more parameters being collected, average follow-up increasing, and more NHS Trusts participating. This dataset offers important opportunities for epidemiological studies and biomedical informatics research, as well as characterising an HBV population for clinical trials through external collaborations with industry.

## Why was the cohort set up?

Chronic infection with hepatitis B virus (HBV) is a global health problem, resulting in an estimated ∼887,000 deaths worldwide in 2015 (1). Unlike deaths from other infections such as tuberculosis, human immunodeficiency virus (HIV) or malaria, the number of viral hepatitis deaths (the majority of which are attributable to HBV and hepatitis C virus (HCV) infection) has increased since 1990 (2). To advance towards international goals for eliminating viral hepatitis (3), it is important to accurately estimate the baseline burden, to develop and deliver interventions based on real-world data, and to monitor progress towards targets at regional and national levels (4).

As the prevalence of HBV infection is low across the United Kingdom (UK) overall, there are limited data describing population characteristics and disease burden (5, 6). Chronic HBV (CHB) nevertheless presents a concern in certain populations, either as a result of increased prevalence, and/or risk factors for the development of long-term liver disease (e.g., chronic coinfection with HIV (7) or other hepatitis viruses (8, 9), diabetes mellitus or metabolic syndrome (10, 11), alcohol abuse (11), migrants from countries/regions with a high prevalence of HBV (12, 13)). Chronic infection can lead to pathology which has a major impact on quality of life and life expectancy, including cirrhosis, end-stage-liver failure, and hepatocellular carcinoma (HCC). Following the successes of direct acting antiviral drugs for HCV treatment as well as potential cure strategies targeting the reservoir in HIV infection, the clinical and research communities have focused progressive attention on cure strategies for HBV. There is therefore a pressing need for national-level data collection to evaluate population characteristics, identify risk factors, assess treatment deployment, develop predictive models for outcomes, and provide a foundation for clinical trials for HBV.

Leveraging existing clinical data is a cost-effective way to build a detailed description of HBV infection. Existing primary care datasets like Clinical Practice Research Datalink (CPRD) (6) do not capture HBV data well, as surveillance and treatment are largely managed in secondary care. During the last decade, large amounts of routinely-collected clinical data have been accumulated in electronic patient record (EPR) systems in the UK’s unified secondary care services. The National Institute for Health Research (NIHR) Health Informatics Collaborative (HIC) collaboration was established in 2014 to enable re-use of these ‘big’ data for translational research (14).

Here we introduce a large prospective multi-centre dataset established through the NIHR HIC viral hepatitis theme collaboration, representing CHB in secondary care across National Health Service (NHS) Trusts (distinct regional organisations, each a separate legal entity, responsible for provision and commissioning of healthcare) in England, UK. The challenges that had to be overcome in order to share data included establishing a unified governance framework across separate organisations, variations in data entry practice, data definitions and clinical practice between sites, de-identification required for large amounts of important free-text data, and different levels of expertise in clinical informatics in different sites (14).

With funding from the NIHR HIC and local support by NIHR Biomedical Research Centres (BRCs) at participating sites, the dataset continues to expand over time, with additional NHS Trusts joining the NIHR HIC viral hepatitis theme, and existing members refining the quality and quantity of data submitted.

## Who is in the cohort?

### Locations and setting

The NIHR HIC HBV cohort is a multisite dataset populated with anonymised routinely-collected clinical data from individuals (including adults and children) with CHB attending secondary care services in 10 NHS Trusts across the UK (**Figure 1**). Current data are from England, but the NIHR HIC provides opportunities to expand the dataset to represent other locations within the UK.

**Figure 1.**
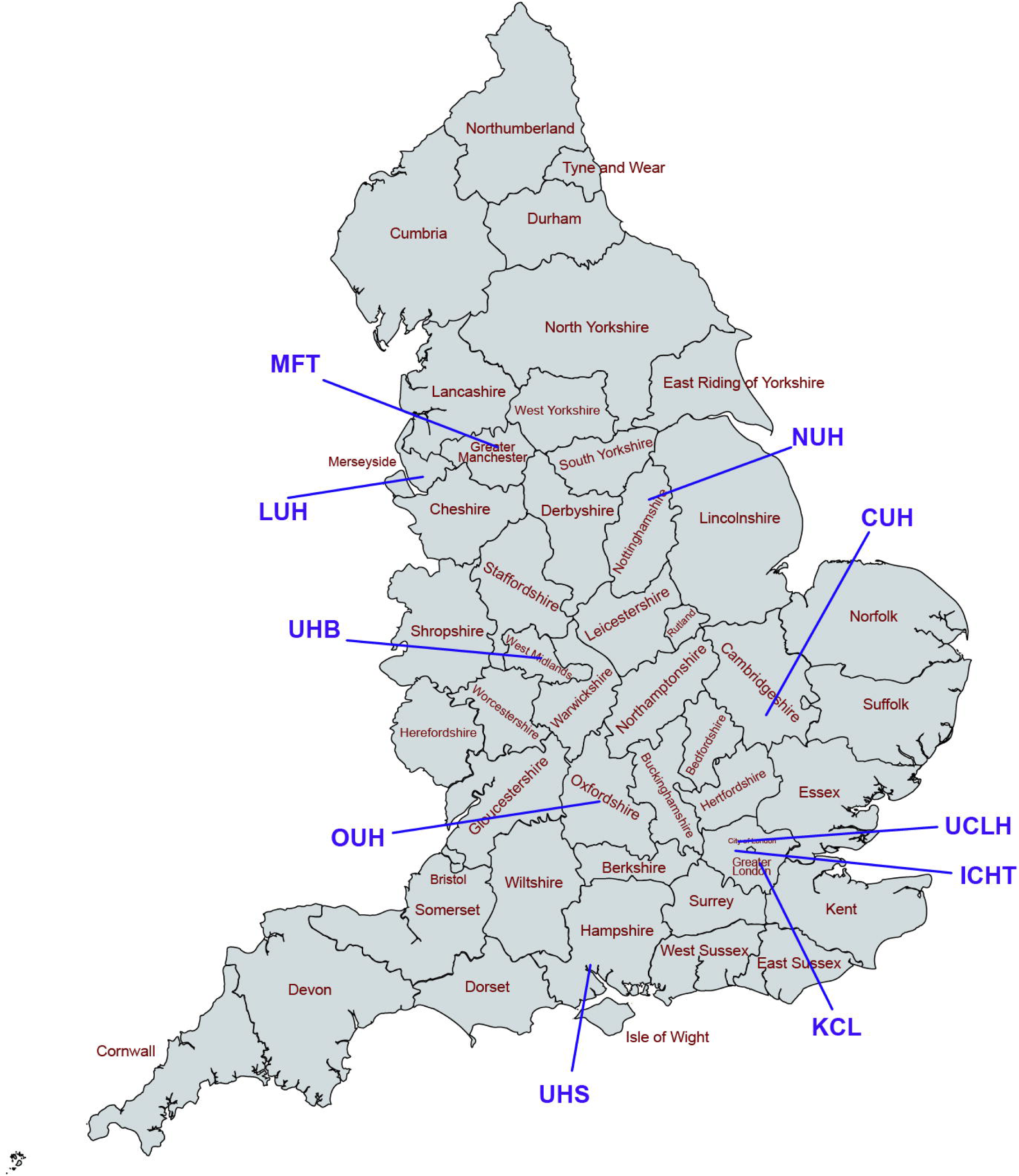
**Locations of the 10 NHS Trusts participating in the NIHR HIC viral hepatitis theme up to Sep 2021**. ***CUH***, *Cambridge University Hospitals NHS Foundation Trust;* ***ICHT***, *Imperial College Healthcare NHS Trust;* ***KCL***, *King’s College Hospital NHS Foundation Trust;* ***LUH***, *Liverpool University Hospitals NHS Foundation Trust;* ***MFT***, *Manchester University NHS Foundation Trust;* ***NUH***, *Nottingham University Hospitals NHS Trust;* ***OUH***, *Oxford University Hospitals NHS Foundation Trust;* ***UCLH***, *University College London Hospitals NHS Foundation Trust;* ***UHB***, *University Hospitals Birmingham NHS Foundation Trust;* ***UHS***, *University Hospital Southampton NHS Foundation Trust*. CUH, ICHT, KCL, OUH, and UCLH were the five NHS Trusts initially included in the NIHR HIC viral hepatitis theme, with LUH, MFT, NUH, UHB, and UHS joining more recently.

At each site, routinely-collected clinical data are captured in local electronic systems. However, these systems were originally designed for hospital operations rather than for research purposes, so data entry practice and storage format are not unified. Different sites use different types of EPR system for clinical solutions (e.g. Cerner Millennium, Epic), and even when sites are using the same type of EPR system, the data record style and integration are locally customised. To overcome such challenges, we have developed an informatics infrastructure and established a comprehensive governance framework for collecting data between heterogenous EPR environments, detailed previously (14). All the laboratory assays used at each site were undertaken on validated platforms in UK labs with clinical accreditation.

Data since the date of EPR system implementation are retrospectively captured, though historical data pre-dating the implementation are also included at some sites. Further data are added prospectively, with updates submitted on request and transferred to the theme central data repository. Thus, the start date (earliest available data) can vary by years between Trusts due to different timelines of EPR system introduction, but the end date (latest available data) is mostly within the same calendar year.

The central data repository is hosted by the theme lead centre (Oxford University Hospitals NHS Foundation Trust) under a governance framework that includes a data sharing agreement and terms on contractual responsibilities, confidentiality, intellectual property, and publication (14). A scientific steering committee, made up of at least one representative from each participating site, meets regularly to review data collection, feedback progress on active projects, consider updates to the database, and review all data requests.

### Eligibility criteria

The inclusion criteria up to May 2021 were (a) individuals for whom data are recorded in the EPR systems; and (b) individuals with CHB, defined by two positive HBsAg tests and/or detectable HBV DNA at least six months apart (**Table S1**). In June 2021, an update was agreed to relax the criteria for inclusion, such that a single positive HBsAg or HBV DNA test was considered sufficient. Although this potentially adds a small number of cases of acute infection, it renders many more cases of chronic infection eligible for data inclusion and thus provides a more complete picture of all HBV infections. The exclusion criteria were: (a) patients without records of demographics, or (b) patients without mandated laboratory data (**Table S2, S3**) in the EPR systems (**Figure S1**).

### Index date, baseline period, and numbers of subjects

For each individual, we defined the first episode of positive HBsAg or HBV DNA recorded in the EPR system as their ‘index date’ (**Figure 2A**). For some patients, the index date may be later than the time when they were clinically diagnosed with CHB due to geographic migration across regions/countries. A baseline period was defined as 365 days within the index date.

**Figure 2.**
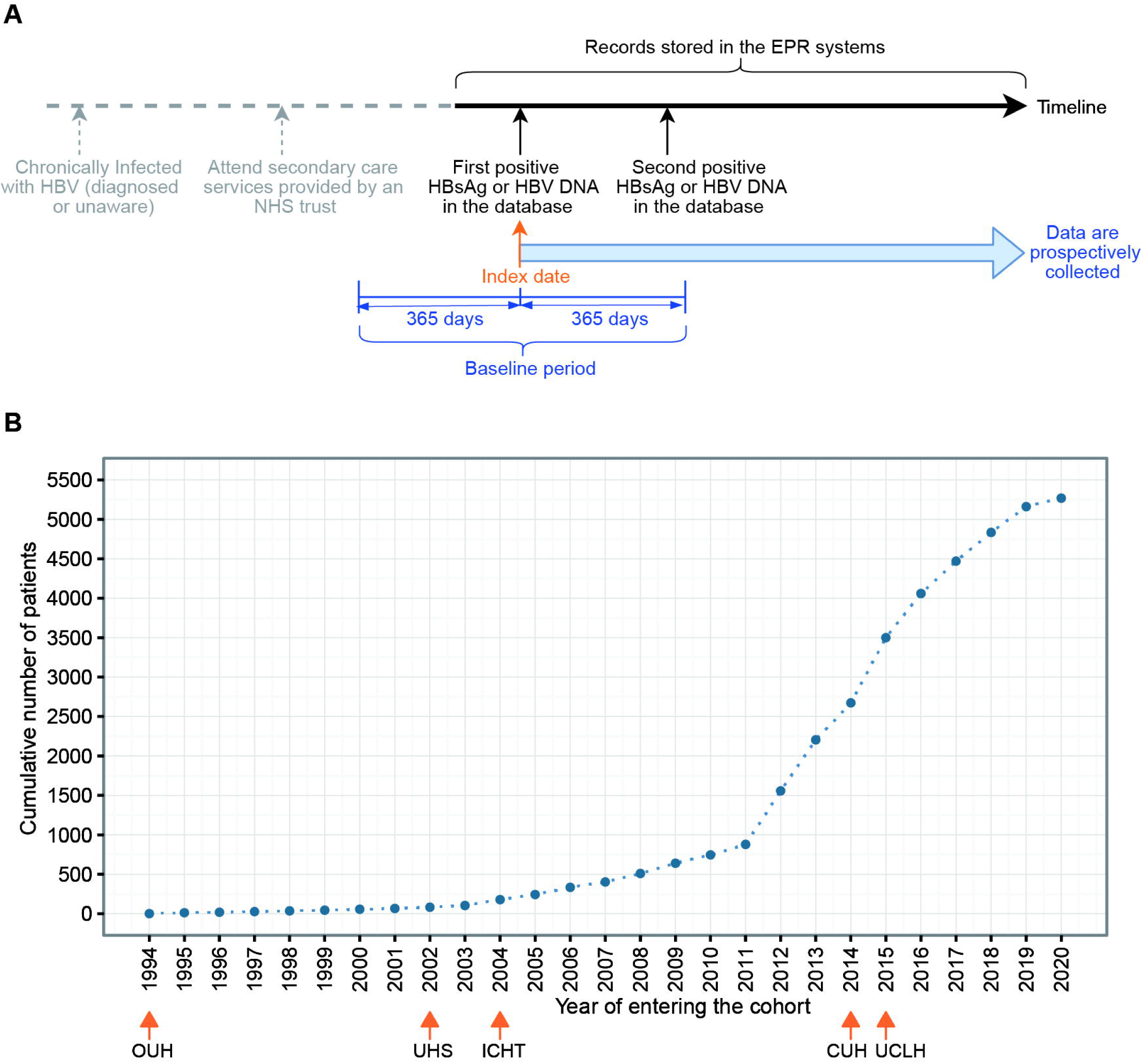
**Index date and number of patients in the NIHR HIC HBV dataset: (A) The timeline and index date definition for an exemplar patient; (B) Cumulative numbers of chronic HBV patients entering the cohort over time.** *In panel A, all data collected after the index date are added into the dataset, and those data before index date are also included if they are available in EPR systems. In panel B, for the five sites which had submitted data, the earliest year of the included data at each site is marked. CUH, Cambridge University Hospitals NHS Foundation Trust; ICHT, Imperial College Healthcare NHS Trust; OUH, Oxford University Hospitals NHS Foundation Trust; UCLH, University College London Hospitals NHS Foundation Trust; UHS, University Hospital Southampton NHS Foundation Trust*.

At the most recent update in August 2021, five of the 10 participating sites submitted data, representing a total of 5269 CHB patients, with index dates between August 1994 and December 2020. Cumulative numbers of cases in the cohort over time are presented in **Figure 2B**. Individuals with age <18 years (n=73) are not described in the remaining text but they are included in the dataset when they reach 18 years of age.

## How often have they been followed up?

Individuals were followed from the index date until they were died or lost to follow-up (defined as no new records within 24 months of the most recent data update). The follow-up frequency of each individual is variable (influenced by clinical requirements and patient preference), and is subject to influence by other factors, including disruptions to clinical services caused by the COVID-19 pandemic since early 2020.

### Follow-up duration and frequency, and availability of longitudinal data

Currently, median follow-up duration of the adult patients in this dataset was 5.0 (IQR: 2.7-7.5) years. 5% (261/5196) of patients died during follow-up, with 9.3 (95% CI: 8.2-10.5) deaths per 1,000 person-years, similar to the mortality rate reported by an Asian study of CHB patients with similar age profile (15). The demographics, follow-up duration, and coinfection characteristics of adults who died (n=261) or were lost to follow-up (n=1071) compared to who are active (n=3864) in the cohort are presented in **Table S4**.

Laboratory parameters such as HBV DNA, HBsAg, alanine aminotransferase (ALT), platelets, and estimated glomerular filtration rate (eGFR) were assessed with a median interval of ∼6 months, and were more frequently measured than hepatitis B virus e antigen (HBeAg), hepatitis B virus e antibody (anti-HBe), and aspartate aminotransferase (AST), assessed with a median interval of 8-10 months (**Figure 3A**). Most patients (87%∼97%) had ≥ 2 ALT, HBV DNA, platelets, and eGFR measurements, whilst a lower proportion (58%, 63%, 73% respectively) had ≥2 HBeAg, anti-HBe, and AST measurements (**Figure 3B**), reflecting differences in clinical practice between sites.

**Figure 3.**
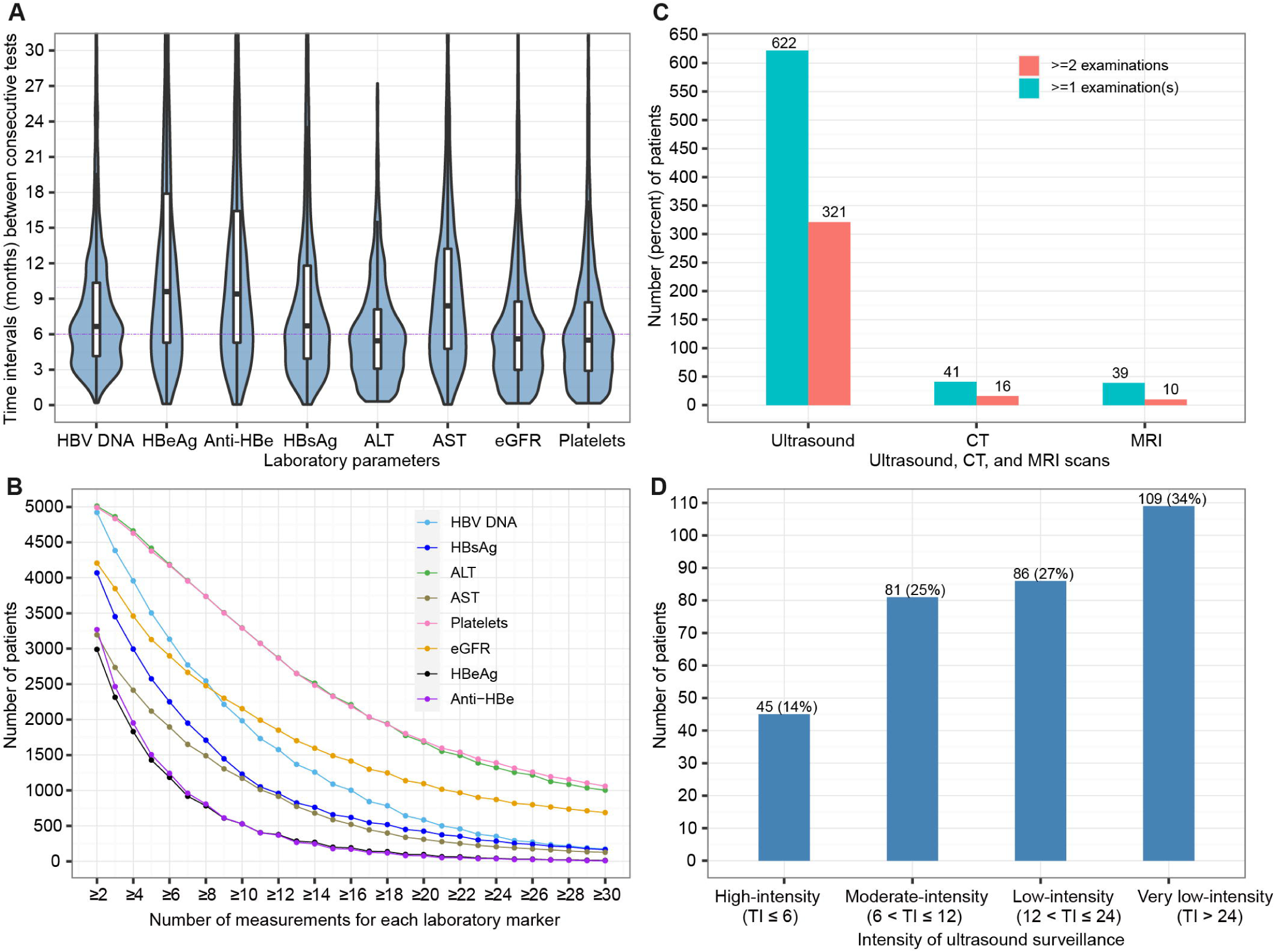
**Follow-up frequency and longitudinal data availability: (A) Time intervals (months) between two consecutive tests within patients of HBV serological and virological biomarkers, liver biochemistry parameters, and renal function markers; (B) Numbers of patients who had longitudinal data (i.e., at least two or more measurements) of laboratory markers (HBV DNA, HBsAg, HBeAg, Anti-HBe, ALT, AST, eGFR, platelets) during the follow-up period; (C) Patients on ultrasound, CT, or MRI surveillance with various numbers of examinations; (D) Patients with various intensity of ultrasound surveillance for those who had ≥ 2 ultrasound scans.** *In panel A, mean value of time intervals between every two consecutive tests within each patient was calculated, then the violin plot and boxplot were drawn based on these mean values with outliers (the observations below the 1st percentile and the observations above the 99th percentile) removed. Boxplots indicate the median and quartiles with whiskers reaching up to 1.5 times the interquartile range. The violin plot outlines illustrate kernel probability density, i.e., the width of the blue shaded area represents the proportion of the data located there. Data beyond 30 months were not shown in the plots. In panel B, numbers under x-axis indicate the number of patients who had longitudinal data with* ≥ *2 measurements on a test. HBV, hepatitis B virus; DNA, deoxyribonucleic acid; HBsAg, hepatitis B virus surface antigen; HBeAg, hepatitis B virus e antigen; Anti-HBe, hepatitis B virus e antibody; ALT, alanine aminotransferase; AST, aspartate aminotransferase; eGFR, estimated glomerular filtration rate; CT, computed tomography; MRI, magnetic resonance imaging; TI, time interval*.

In line with clinical guidelines, ultrasound is routinely used for surveillance; CT and MRI scans are less frequently used, if concerns are raised by other imaging or clinical biomarkers. Two sites (contributing data for 1087 adults with CHB) have submitted imaging reports. During follow-up, 622/1087 (57.2%) and 321/1087 (29.5%) patients had ≥1 and ≥2 ultrasound examination(s) respectively (**Figure 3C**). For those with ≥ 2 ultrasound examinations, 14% were on high-intensity surveillance (≤6 months), 27% on moderate-intensity surveillance (>6-12 months), 25% on low-intensity surveillance (>12-24 months), and 34% on surveillance with intervals >24 months (**Figure 3D**).

## What has been measured?

### Data model

A standardised data model, used by all collaborating sites for data mapping, extraction, and submission, has been designed and released in the Mauro Data Mapper (used as the NIHR HIC’s metadata catalogue, https://modelcatalogue.cs.ox.ac.uk/nihr-hic/#/home). An overview of data classes and elements defined in the data model is provided in **Table S2**. Death records are collected from the Office of National Statistics (ONS) through NHS Digital’s Spine portal, and date of death is checked for each patient before data are submitted. All data classes are linked to produce a complete record for each unique patient. Data are then anonymised before submission, with each unique patient assigned with a study identifier; this allows researchers to conduct analysis at the individual level without the possibility of patient identification.

### Data inference

One principle of the designed data model is to collect source data as it appears in EPR systems and to allow researchers to infer information of interest using raw data collected. All the inferred fields included in the data model are presented in the supplementary XML schema definition (XSD) file. Here, we used the inferred variables coinfection exposures and liver disease severity.

Chronic viral co-infections (HIV, HCV, HDV), and acute infection or past exposure to hepatitis E virus (HEV), were identified from laboratory tests (**Table S3**). Liver fibrosis and cirrhosis were characterised based on Ishak or METAVIR scores from biopsy reports (16) or liver stiffness measurements from transient elastography (FibroScan) if available; otherwise, we used AST to platelet ratio index (APRI) (17) or Fibrosis-4 (FIB-4) scores (18) (**Figure S2)**. We used pre-defined thresholds for significant/advanced fibrosis and cirrhosis: 1.5 and 2.0 for APRI score respectively (17); 3.25 and 3.6 for FIB-4 score respectively (18, 19). Decompensation and HCC information was retrieved from clinical and imaging reports if available.

## What has it found?

### Baseline characteristics

#### Demographic, HBV serological and virological characteristics, coinfections

At baseline, for adults (n = 5196), the median age was 40 years (IQR: 32-50) and 55% were male; 4143 had ethnicity recorded, among whom 30% were Asian, 24% were Black, 30% were White, and the remaining 16% were mixed or other ethnic groups (**Table 1)**.

**Table 1.**
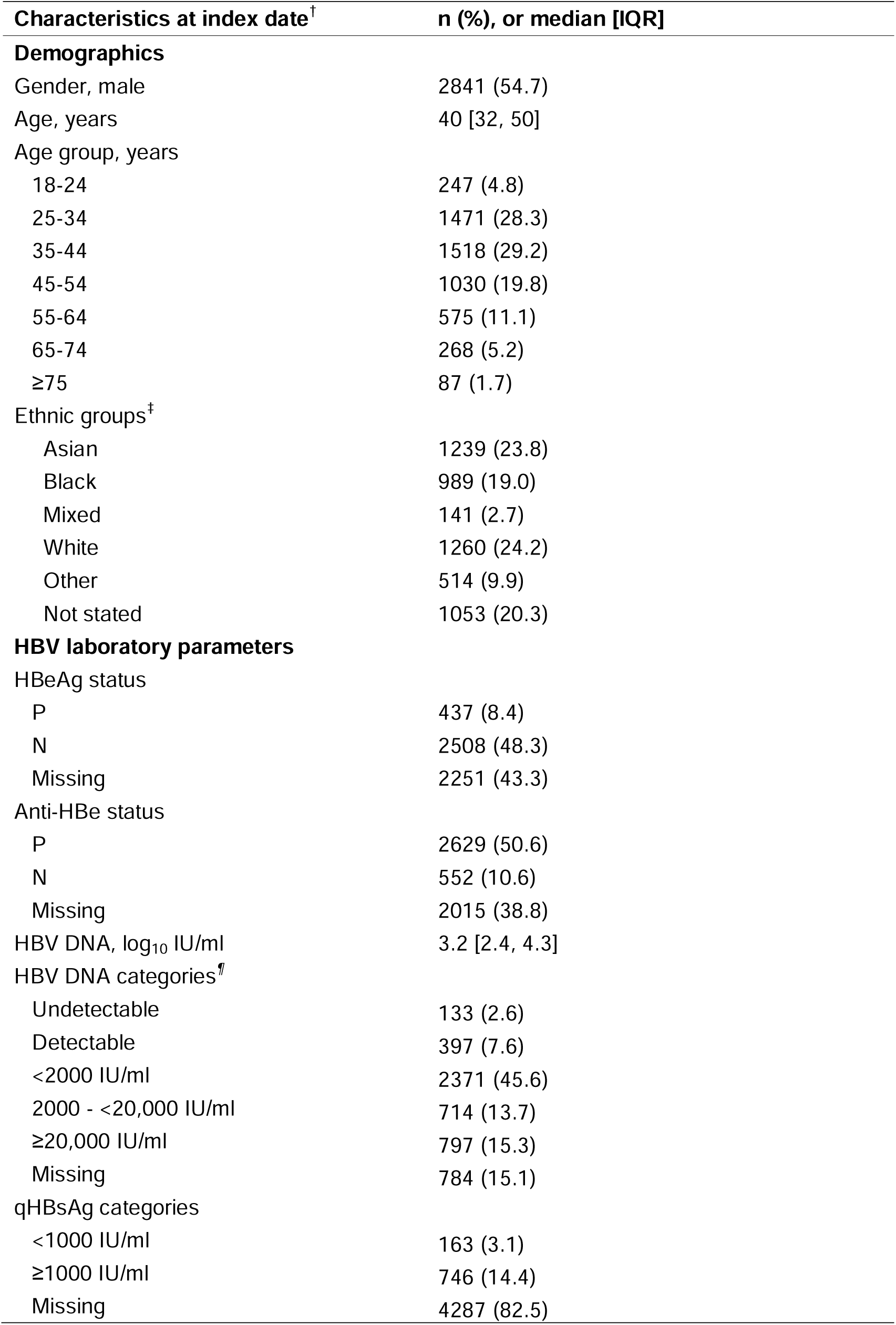
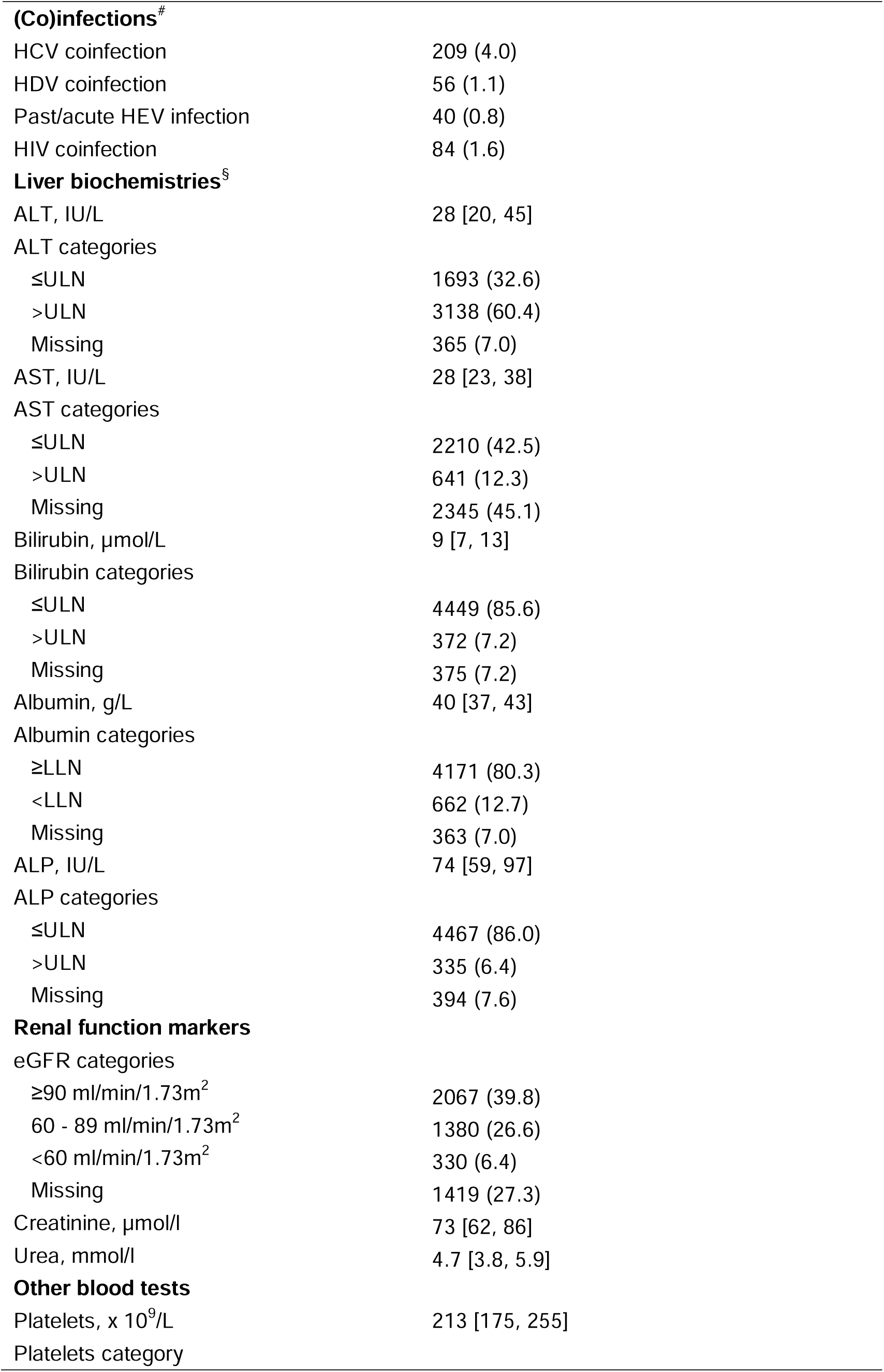
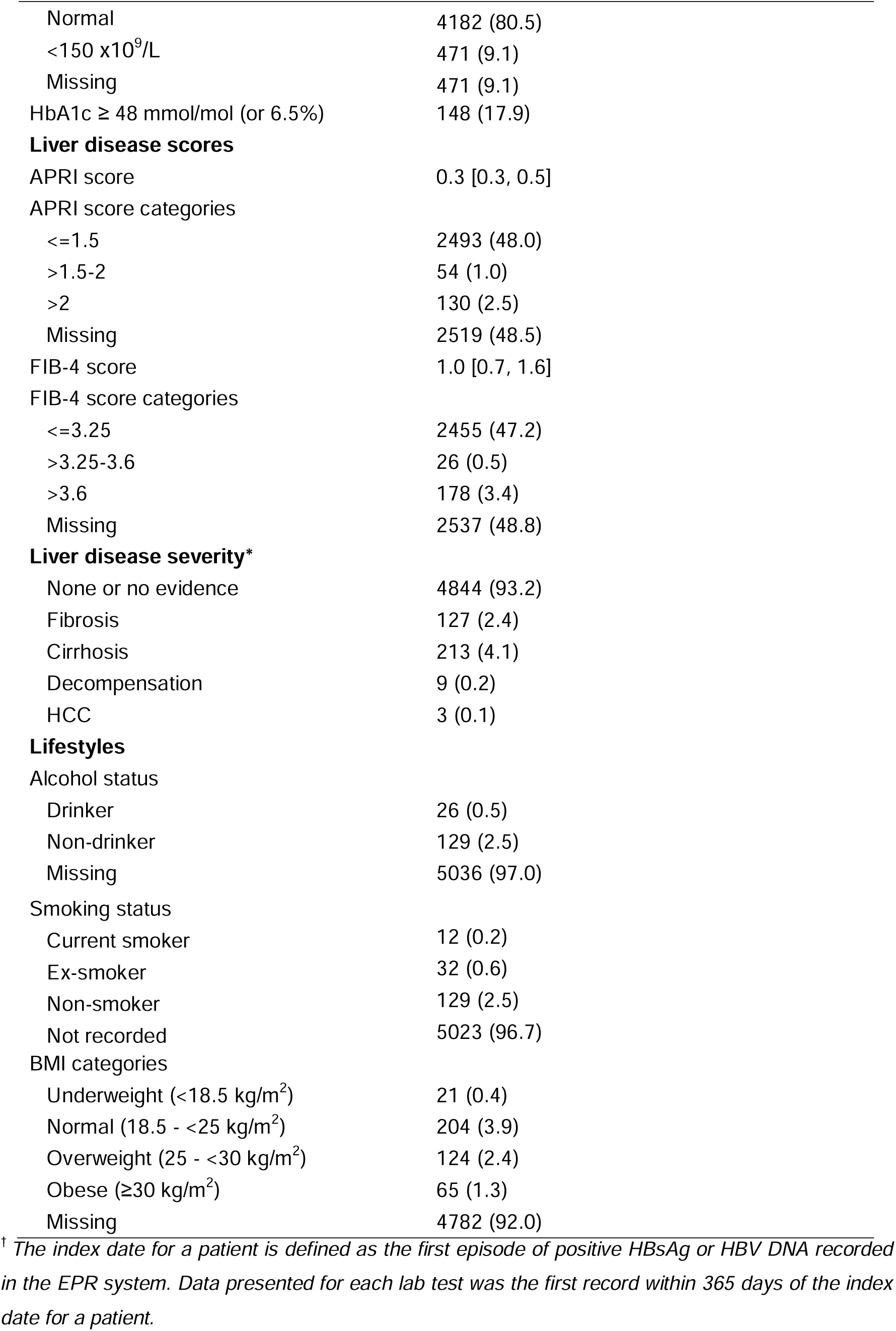
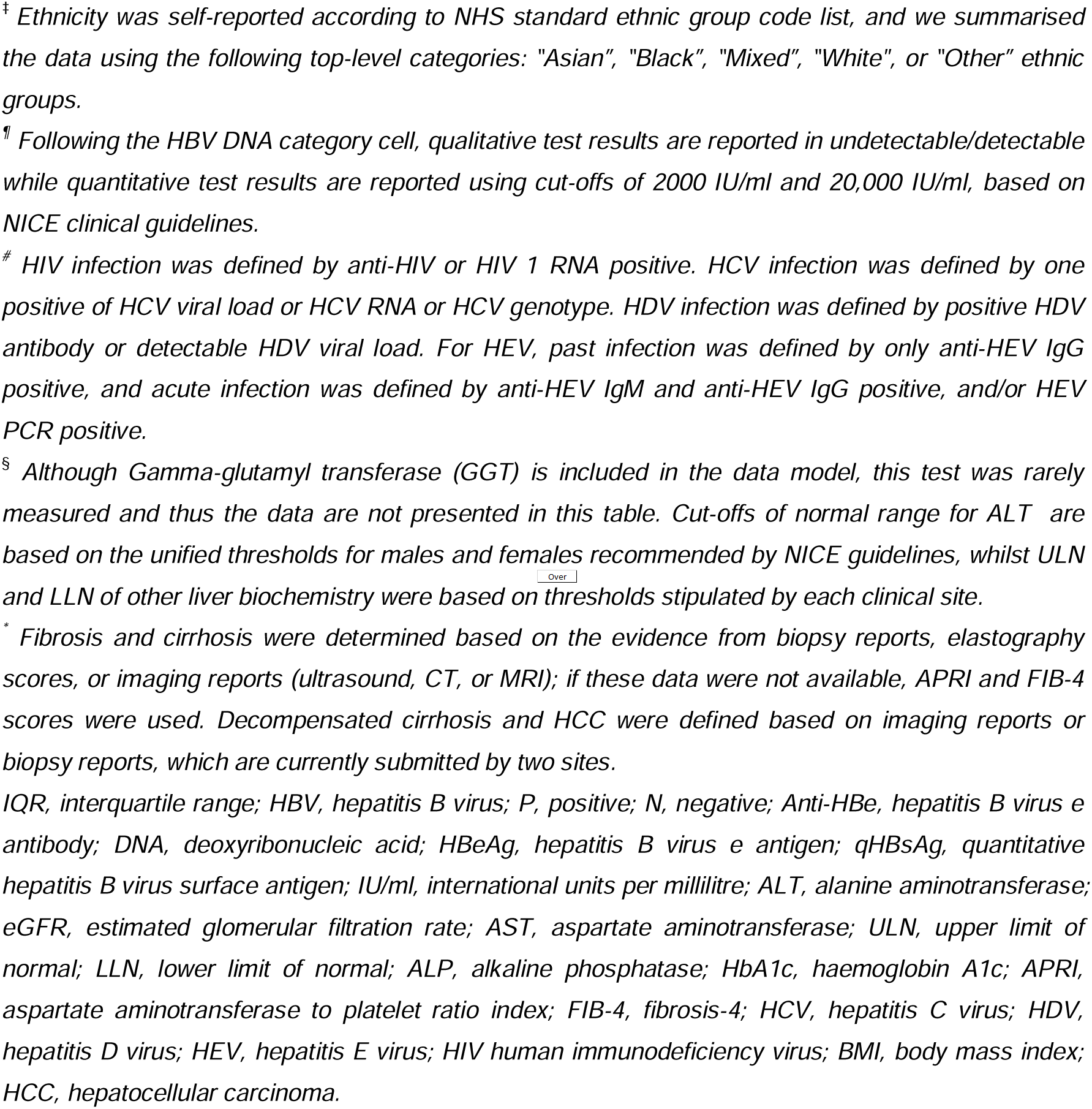
Demographic, serological and virological characteristics of individuals with chronic HBV infection collected from five secondary care centres in England (baseline data, n=5196).

| **Characteristics at index date<sup>†</sup>** | **n (%), or median [IQR]** |
| --- | --- |
| **Demographics** |  |
| Gender, male | 2841 (54.7) |
| Age, years | 40 [32, 50] |
| Age group, years |  |
| 18-24 | 247 (4.8) |
| 25-34 | 1471 (28.3) |
| 35-44 | 1518 (29.2) |
| 45-54 | 1030 (19.8) |
| 55-64 | 575 (11.1) |
| 65-74 | 268 (5.2) |
| ≥75 | 87 (1.7) |
| Ethnic groups<sup>‡</sup> |  |
| Asian | 1239 (23.8) |
| Black | 989 (19.0) |
| Mixed | 141 (2.7) |
| White | 1260 (24.2) |
| Other | 514 (9.9) |
| Not stated | 1053 (20.3) |
| **HBV laboratory parameters** |  |
| HBeAg status |  |
| P | 437 (8.4) |
| N | 2508 (48.3) |
| Missing | 2251 (43.3) |
| Anti-HBe status |  |
| P | 2629 (50.6) |
| N | 552 (10.6) |
| Missing | 2015 (38.8) |
| HBV DNA, log<sub>10</sub> IU/ml | 3.2 [2.4, 4.3] |
| HBV DNA categories<sup>¶</sup> |  |
| Undetectable | 133 (2.6) |
| Detectable | 397 (7.6) |
| <2000 IU/ml | 2371 (45.6) |
| 2000 - <20,000 IU/ml | 714 (13.7) |
| ≥20,000 IU/ml | 797 (15.3) |
| Missing | 784 (15.1) |
| qHBsAg categories |  |
| <1000 IU/ml | 163 (3.1) |
| ≥1000 IU/ml | 746 (14.4) |
| Missing | 4287 (82.5) |

| Characteristics at index date<sup>†</sup> | n (%), or median [IQR] |
| --- | --- |
| **(Co)infections<sup>#</sup>** |  |
| HCV coinfection | 209 (4.0) |
| HDV coinfection | 56 (1.1) |
| Past/acute HEV infection | 40 (0.8) |
| HIV coinfection | 84 (1.6) |
| **Liver biochemistries<sup>§</sup>** |  |
| ALT, IU/L | 28 [20, 45] |
| ALT categories |  |
| ≤ULN | 1693 (32.6) |
| >ULN | 3138 (60.4) |
| Missing | 365 (7.0) |
| AST, IU/L | 28 [23, 38] |
| AST categories |  |
| ≤ULN | 2210 (42.5) |
| >ULN | 641 (12.3) |
| Missing | 2345 (45.1) |
| Bilirubin, µmol/L | 9 [7, 13] |
| Bilirubin categories |  |
| ≤ULN | 4449 (85.6) |
| >ULN | 372 (7.2) |
| Missing | 375 (7.2) |
| Albumin, g/L | 40 [37, 43] |
| Albumin categories |  |
| ≥LLN | 4171 (80.3) |
| <LLN | 662 (12.7) |
| Missing | 363 (7.0) |
| ALP, IU/L | 74 [59, 97] |
| ALP categories |  |
| ≤ULN | 4467 (86.0) |
| >ULN | 335 (6.4) |
| Missing | 394 (7.6) |
| **Renal function markers** |  |
| eGFR categories |  |
| ≥90 ml/min/1.73m<sup>2</sup> | 2067 (39.8) |
| 60 - 89 ml/min/1.73m<sup>2</sup> | 1380 (26.6) |
| <60 ml/min/1.73m<sup>2</sup> | 330 (6.4) |
| Missing | 1419 (27.3) |
| Creatinine, µmol/l | 73 [62, 86] |
| Urea, mmol/l | 4.7 [3.8, 5.9] |
| **Other blood tests** |  |
| Platelets, x 10<sup>9</sup>/L | 213 [175, 255] |
| Platelets category |  |
| Normal | 4182 (80.5) |
| <150 x10<sup>9</sup>/L | 471 (9.1) |
| Missing | 471 (9.1) |
| HbA1c ≥ 48 mmol/mol (or 6.5%) | 148 (17.9) |
| **Liver disease scores** |  |
| APRI score | 0.3 [0.3, 0.5] |
| APRI score categories |  |
| ≤1.5 | 2493 (48.0) |
| >1.5-2 | 54 (1.0) |
| >2 | 130 (2.5) |
| Missing | 2519 (48.5) |
| FIB-4 score | 1.0 [0.7, 1.6] |
| FIB-4 score categories |  |
| ≤3.25 | 2455 (47.2) |
| >3.25-3.6 | 26 (0.5) |
| >3.6 | 178 (3.4) |
| Missing | 2537 (48.8) |
| **Liver disease severity\*** |  |
| None or no evidence | 4844 (93.2) |
| Fibrosis | 127 (2.4) |
| Cirrhosis | 213 (4.1) |
| Decompensation | 9 (0.2) |
| HCC | 3 (0.1) |
| **Lifestyles** |  |
| Alcohol status |  |
| Drinker | 26 (0.5) |
| Non-drinker | 129 (2.5) |
| Missing | 5036 (97.0) |
| Smoking status |  |
| Current smoker | 12 (0.2) |
| Ex-smoker | 32 (0.6) |
| Non-smoker | 129 (2.5) |
| Not recorded | 5023 (96.7) |
| BMI categories |  |
| Underweight (<18.5 kg/m<sup>2</sup>) | 21 (0.4) |
| Normal (18.5 - <25 kg/m<sup>2</sup>) | 204 (3.9) |
| Overweight (25 - <30 kg/m<sup>2</sup>) | 124 (2.4) |
| Obese (≥30 kg/m<sup>2</sup>) | 65 (1.3) |
| Missing | 4782 (92.0) |

<sup>†</sup> The index date for a patient is defined as the first episode of positive HBsAg or HBV DNA recorded in the EPR system. Data presented for each lab test was the first record within 365 days of the index date for a patient.
<sup>‡</sup> Ethnicity was self-reported according to NHS standard ethnic group code list, and we summarised the data using the following top-level categories: “Asian”, “Black”, “Mixed”, “White”, or “Other” ethnic groups.
<sup>¶</sup> Following the HBV DNA category cell, qualitative test results are reported in undetectable/detectable while quantitative test results are reported using cut-offs of 2000 IU/ml and 20,000 IU/ml, based on NICE clinical guidelines.
<sup>#</sup> HIV infection was defined by anti-HIV or HIV 1 RNA positive. HCV infection was defined by one positive of HCV viral load or HCV RNA or HCV genotype. HDV infection was defined by positive HDV antibody or detectable HDV viral load. For HEV, past infection was defined by only anti-HEV IgG positive, and acute infection was defined by anti-HEV IgM and anti-HEV IgG positive, and/or HEV PCR positive.
<sup>§</sup> Although Gamma-glutamyl transferase (GGT) is included in the data model, this test was rarely measured and thus the data are not presented in this table. Cut-offs of normal range for ALT are based on the unified thresholds for males and females recommended by NICE guidelines, whilst ULN and LLN of other liver biochemistry were based on thresholds stipulated by each clinical site.
<sup>\*</sup> Fibrosis and cirrhosis were determined based on the evidence from biopsy reports, elastography scores, or imaging reports (ultrasound, CT, or MRI); if these data were not available, APRI and FIB-4 scores were used. Decompensated cirrhosis and HCC were defined based on imaging reports or biopsy reports, which are currently submitted by two sites.
IQR, interquartile range; HBV, hepatitis B virus; P, positive; N, negative; Anti-HBe, hepatitis B virus e antibody; DNA, deoxyribonucleic acid; HBeAg, hepatitis B virus e antigen; qHBsAg, quantitative hepatitis B virus surface antigen; IU/ml, international units per millilitre; ALT, alanine aminotransferase; eGFR, estimated glomerular filtration rate; AST, aspartate aminotransferase; ULN, upper limit of normal; LLN, lower limit of normal; ALP, alkaline phosphatase; HbA1c, haemoglobin A1c; APRI, aspartate aminotransferase to platelet ratio index; FIB-4, fibrosis-4; HCV, hepatitis C virus; HDV, hepatitis D virus; HEV, hepatitis E virus; HIV human immunodeficiency virus; BMI, body mass index; HCC, hepatocellular carcinoma.

Baseline HBeAg and anti-HBe status were available for 2945 and 3181 patients respectively. 437/2945 (15%) were HBeAg-positive and 83% (2629/3181) were anti-HBe positive. 4412/5196 (85%) patients had HBV DNA tests at baseline. For those with quantitative results (n=3882), the median HBV DNA viral load was 3.2 (IQR: 2.4-4.3) log_10_ IU/ml; 18% had moderate-high levels (2000 - <20,000 IU/ml) and 21% had high levels (≥ 20,000 IU/ml), based on thresholds recommended by National Institute for Health and Care Excellence (NICE) clinical guidelines (20). Quantitative HBsAg is not routinely tested at most sites, with baseline data available for just 909 individuals. Among these, 18% had low HBsAg levels (<1000 IU/ml) whilst 82% had high HBsAg levels (≥1000 IU/ml), based on the NICE threshold (20).

At baseline, 4.0%, 1.1%, and 1.6% were coinfected with HCV or HDV or HIV, respectively, and a smaller proportion coinfected with past/acute HEV (n=40; 0.8%).

#### Liver biochemistry, renal function markers and other laboratory characteristics at baseline

The most frequently tested liver enzyme is ALT, available for 93% (4831/5196) of patients, among whom 65% (3138/4831; 64% for males and 66% for females) had elevated levels (≥30 IU/L for males, ≥19 IU/L for females as per NICE guidelines). AST is not routinely measured at some sites, but available for a subset of individuals (n=2851), among whom 22% (641/2851) had elevated levels (set by sites’ laboratories). Data for other routine liver biochemistry including albumin, bilirubin, and alkaline phosphatase were available for 92.4%-93.0% of patients ; but these three tests were less likely to be deranged in CHB patients (14%, 8%, and 7%, respectively), compared to ALT and AST.

For those with eGFR available (n=3777), 37% (1380/3777) had mildly decreased renal function (eGFR 60-89 ml/min/1.73m^2^), and 9% (330/3777) had moderate/severe impairment of renal function (eGFR <60 ml/min/1.73m^2^). Median platelet count (measured for 91% of patients) was 213 x10^9^/L (IQR: 175-255), with thrombocytopenia (<150 x10^9^/L) in 9%. HbA1c was not frequently tested in this sample of CHB patients, but among those with HbA1c (n=825) available, 18% had ≥48 mmol/mol (or 6.5%), which might indicate diabetes as per NICE thresholds (21).

#### Liver disease severity at baseline

To evaluate liver disease severity, we examined laboratory tests, elastography, imaging (ultrasound, MRI, CT) reports, and biopsy reports. AST, ALT and platelets (≤12 weeks apart) were used to calculate APRI and FIB-4 scores, resulting in these two scores both available in ∼51% of cases (2677/5196 and 2659/5196 respectively) (**Table 1**). Among these, 6.9% (184/2677) had APRI score >1.5 and 7.7% (204/2659) had FIB-4 score >3.25 (17-19). To date, elastography data have not been routinely recorded by EPR systems in a way that allows capture by an automated data pipeline, and are thus currently available for a small minority of individuals. Ongoing work is being undertaken to collate this information. For the two sites that have so far submitted imaging or biopsy reports, 378/1087 (35%) patients had these reports available at baseline; a diagnosis of decompensation and HCC was suggested in 9/378 (2.4%) and 3/378 (0.8%), respectively.

With combining information from imaging/biopsy reports, elastography scores, or APRI and FIB-4 scores (**Figure S2**), 2.4%, 4.1% and 0.3% had fibrosis, cirrhosis, and decompensation/HCC at baseline respectively (**Table 1**). However, these hepatic complications may currently be underestimated at baseline due to missing data, and may increase over time, as we have previously quantified (22).

#### Lifestyle data

Alcohol, smoking, and BMI data were available for a small proportion of cases at baseline, as these data have not been consistently captured by EPR systems, but as data mining and recording improves these parameters will be prospectively added.

### Antiviral treatment

Among patients with available treatment information (n=4421), 19% (828/4421) received HBV treatment at any time during the data collection period. Follow-up duration was similar between treated and untreated patients (**Table S5**). Patients who received treatment were more likely to be male, older at baseline, and more likely to be Asian than from other ethnic groups (all P<0.001, **Table S5, Figure 4A-B**). Reflecting UK HBV management guidelines (20), tenofovir disoproxil fumarate (TDF) was the most frequently prescribed antiviral treatment (61%; 505/828), followed by entecavir (ETV, 19%; 161/828) (**Figure 4C**). With further stratification by coinfections, patients with HIV, HCV, HDV, or HEV coinfections accounted for 44/505 (9%) in the group prescribed TDF (**Figure S3**).

**Figure 4.**
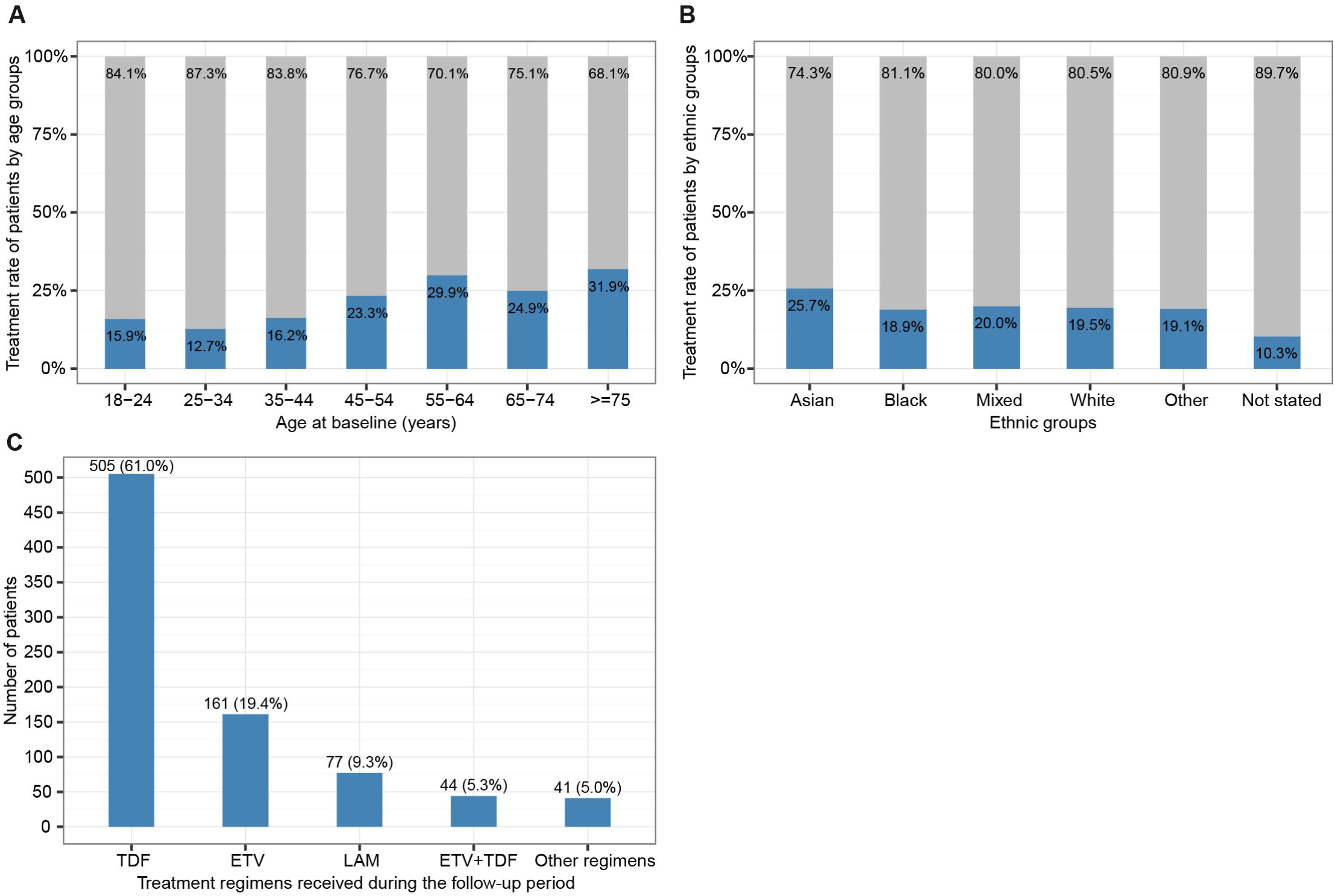
**Percentage of patients receiving antiviral treatment for chronic HBV infection in different (A) age groups and (B) ethnic groups; (C) Treatment regimens received by patients who were on treatment during the follow-up period.** *In panel A, In panel B, ethnic groups were based on the NHS ethnic categories. In panel C, those regimens infrequently prescribed for patients are shown together in the final column, such as combinations of LAM+TDF, ETV+LAM, ADV+LAM, or ETV+LAM+TDF. There was not any episode of interferon drug recorded for the cohort although this drug is included in the data model. Each percent value in panel C was calculated as the number of patients receiving a type of regimen divided by the total number of patients who were on treatment. TDF, tenofovir disoproxil fumarate; ETV, entecavir; LAM, Lamivudine; ADV, adefovir dipivoxil. Treatment data are currently available from four sites*.

### Key findings

This present cohort comprises CHB patients of diverse ethnicities from five secondary care NHS Trusts across England, mostly representing adults in middle life. The proportion of patients receiving antiviral treatment varies by gender, age, and ethnicity, which warrants further investigation. A large majority of patients in this cohort had longitudinal measurements of relevant laboratory parameters, providing promising opportunities for longitudinal analyses.

We have already undertaken studies using this framework, with more in process. During COVID-19 pandemic, we have investigated service disruptions, revealing that reduction in rates of surveillance closely track COVID-19 incidence and periods of population lock-down. Using this dataset, we have reported a bimodal viral load distribution (23), and found evidence of a virological set point in untreated patients (22, 23). In a comparison of TDF-treated vs. untreated patients, we reported variable ethnicity distributions across the two groups, and some evidence for liver fibrosis progression in the untreated group, highlighting a need for further evidence for expanded treatment (22). A study of HBsAg and HBeAg clearance dynamics demonstrated that these markers may contribute to prognostication and patient-stratified care, and provide a foundation for advancing insights into mechanisms of disease control (24). The list of publications is available in https://hic.nihr.ac.uk, with ongoing/planned studies presented in **Table S6**.

## What are the main strengths and weaknesses?

As the biggest dataset reflecting CHB in secondary care in England, and growing year on year with improving quality, NIHR HIC HBV dataset is an invaluable resource for answering diverse questions, supporting collaborations, refining approaches for care stratification and treatment, and influencing policy for health interventions. As the HBV field moves towards new therapeutics, with a quest for cure strategies, clear information about the characteristics of HBV infection in different settings will be essential to underpin the design and implementation of clinical trials, and ultimately to inform equitable access to treatment.

The strengths of NIHR HIC HBV derive from the broad interdisciplinary and cross-site collaboration among clinicians (including hepatology, infectious disease and microbiology specialists), informaticians, project managers, and data managers/analysts, representing the NHS Trusts, the NIHR BRCs, and the affiliated universities actively participating in the research. Each NHS Trust publishes regularly updated Patient and Public Involvement strategies and engages with patients and the public about the research supported using Trust resources on a regular basis.

The multi-site approach integrates CHB data for a broad cross-section of populations from secondary care, and produces comprehensive records for each individual, with an automatic data validation process. Longitudinal clinical data are particularly important for informing treatment and stratification. Data collection is continuing, with the sample size growing, collection of more parameters being completed, average follow-up increasing, and expanding to include more NHS Trusts across the UK. The diversity and statistical power of the dataset will be therefore enhanced for future analyses, providing robust and reliable results despite heterogeneous intrinsic characteristics exist in patients from different sources.

We recognise limitations which can influence data quality and completeness. Although assays are performed on validated platforms, methods of laboratory tests vary by site or period, e.g., variable equations are used for eGFR calculation. Therefore, data may need calibration or transformation before analyses or must be flagged during data comparison. As different Trusts prioritise different tests, various levels of missingness exist in liver biochemistry like AST, and serology markers such as HBeAg and anti-HBe; though these data can influence planning and improving the standard of care as well as access of patients to new treatments aiming at immunological control. Data at some sites are not currently linked to national registries/sources such as ONS death registrations. Additionally, free-text imaging and liver biopsy reports are not systematically available as anonymisation processes that are novel to some sites must be performed before data can be shared. Meanwhile, some data points are difficult to capture from EPR systems, such as treatment records stored in local pharmacy systems, elastography scores recorded in inconsistent and inaccessible formats, and self-reported alcohol data not consistently recorded. Although data noise is a common limitation accompanying use of routinely collected clinical data, findings will become more robust as larger study populations are assimilated, electronic systems become better at data capture, and the data model is further refined.

Our original inclusion criteria required two episodes of positive HBsAg and/or HBV DNA tests ≥ 6 months apart, which might result in some cases with missing data being excluded. The relaxation of the inclusion criteria from June 2021 to one positive HBsAg and/or HBV DNA test will provide a wider population available for investigation, but allowing researchers to apply their own criteria to narrow down the population to include only the more stringent diagnosis of CHB if required for a particular question. Additionally, many individuals with HBV infection are not diagnosed or not receiving clinical care, and thus not represented in secondary care datasets. These individuals may include a disproportionate number in vulnerable groups, including migrants (25) (and perhaps specifically non-English speakers), people who inject drugs (26), and those in prison or detention centres (27).

Although comparable HBV datasets are more available in other countries, such as China and the US (28-30), there are scarce comprehensive data of HBV in the UK, except data reported from certain populations (31-33) or the primary care population (5, 6). We believe this secondary care cohort can start to fill evidence gaps, especially by collating laboratory and imaging data, which are not well captured in primary care.

As an exemplar case, this cohort profile not only highlights the potential utility of a CHB cohort, but also demonstrates that routine clinical data is a valuable resource for translational research. Our use of data during the COVID-19 pandemic (34) highlights how the resource can be quickly adapted to address new questions as they arise.

## Can I get hold of the data? Where can I find out more?

Any potential collaborations are welcomed, and data are available to researchers on request following review by the steering committee. Further details are available at https://hic.nihr.ac.uk. Queries regarding data access should be directed to.

## Supporting information

Supplementary XSD file (data model)

## Data Availability

Any potential collaborations are welcomed, and data are available to researchers on request following review by the steering committee of NIHR HIC viral hepatitis theme. Further details are available at https://hic.nihr.ac.uk. Queries regarding data access should be directed to.

## Declarations

### Ethics approval

The research database for the NIHR HIC viral hepatitis theme was approved by South Central - Oxford C Research Ethics Committee (REF Number: 21/SC/0060). All methods in this study were carried out in accordance with relevant guidelines and regulations.

### Supplementary data

Supplementary data are available online.

### Author contributions

EB, PCM, GSC, WG, EN, SIK, KW, and JD contributed to the conception or design of the work. EB, PCM, JD, KW, GSC, EN, WG, and SIK directed the study’s implementation. TW, DAS, CC, OF, ZM, TN, SH, HS, KAV, GR, SL, BG, LM, DP, CRJ, VT, AC, HP, FB, JO, FW, LR, DR, LE, PK, MA, and JC contributed to the methodology development and the acquisition, processing, interpretation, or management of study data. TW, DAS, CC designed the analytical strategy and TW conducted the analysis, supervised by EB and PCM. PCM, EB, WG, EN, GSC, SIK, PK, MA, and JC helped interpret the findings. TW, PCM, EB, DAS, CC, and KAV contributed to the original draft. All authors reviewed and revised the manuscript critically and approved the final version for publication.

## Acknowledgments

The authors would like to thank all the research nurses and research admin staff at the contributing sites for their help in data collection and submission. The authors would also like to thank clinicians, projects managers, informaticians, and data managers at the other participating sites (Nottingham University Hospitals NHS Trust; Manchester University NHS Foundation Trust; Liverpool University Hospitals NHS Foundation Trust; University Hospitals Birmingham NHS Foundation Trust; King’s College Hospital NHS Foundation Trust) working to submit data.

