## Supplementary Tables and Figures for "Cohort Profile: National Institute for Health Research Health Informatics Collaborative: Hepatitis B Virus (NIHR HIC HBV) Research Dataset"

^#^ Co-last authors

* Corresponding authors. The Peter Medawar Building for Pathogen Research, South Parks Road, Oxford, OX1 3SY, UK.

### Supplementary Tables and Figures

Table S1. The possible combinations of tests for identifying patients with chronic HBV infection up to June 2021*

| **First Test** | **Second Test** | **Time interval** |
| --- | --- | --- |
| HBsAg Positive | HBsAg Positive | ≥6 months |
| HBsAg Positive | HBV DNA Detectable | ≥6 months |
| HBV DNA Detectable | HBsAg Positive | ≥6 months |
| HBV DNA Detectable | HBV DNA Detectable | ≥6 months |

HBsAg, hepatitis B virus surface antigen; HBV, hepatitis B virus; DNA, deoxyribonucleic acid.

** These criteria apply to the whole dataset described here. Entry criteria were relaxed at this date to provide a more inclusive approach to collection of all HBV data, such that new individuals identified prospectively will only require a single HBV DNA or HBsAg positive result to be included.*

Table S2. Data classes and data elements defined in the data model *

| **Data class** | **Data elements** | **Priority** | **Version 2.0.0** | **Version 2.1.0** |
| --- | --- | --- | --- | --- |
| Demographics^#^ | Year of birth, gender, ethnic code, country of birth | Mandatory | √ | √ |
| Laboratory tests^§^ | Test name, date time, result, result numerical, units, lower reference value, upper reference value, point of care test | Mandatory | √ | √ |
| Deaths^#^ | Year of death, reason for death | Desirable | √ | √ |
| HBV treatments | Regimen, start date, end date | Desirable | √ | √ |
| HCV treatments | Regimen, start date, end date, inferred SVR12, reported SVR12 | Desirable | √ | √ |
| Elastography | Date, liver stiffness, method, units | Desirable | √ | √ |
| Ultrasound reports | Date, report | Desirable | √ | √ |
| CT scan reports | Date, report | Desirable | √ | √ |
| MRI scan reports | Date, report | Desirable | √ | √ |
| Biopsy reports | Date, report | Desirable | √ | √ |
| Liver disease progression^†^ | Date recorded, method, name, value, value numeric, site provided^¶^ | Desirable | √ | √ |
| Hepatitis diagnoses | Date of first positive test, hepatitis virus, site provided^¶^ | Desirable | √ | √ |
| Risk factors^‡^ | Date recorded, inference method, name value, value numeric, units, site provided^¶^ | Optional | √ | √ |
| ICD diagnosis | Diagnosis date, ICD code, code order, diagnosis description | Desirable |  | √ |
| Deprivation score | IMDD, year postcode recorded | Desirable |  | √ |
| Healthcare utilisation | Nature of attendance, date of arrival, reason for attendance, date of departure | Desirable |  | √ |

** Detailed definitions of data classes and data elements are provided in the supplementary* XSD file. *Ideally, data are submitted in a defined Extensible Markup Language (XML) format, and then automatically validated and loaded to the central data repository of NIHR HIC viral hepatitis theme. If data are not submitted in XML format, they are transformed by the theme lead centre and then checked by the submitting site. Three new data classes (ICD codes, deprivation scores, health utilisation) along with their corresponding data elements have been added in the version 2.1.0 of the data model in Sep 2021.*

^#^ *Definitions for data elements, including ethnicity, country of birth, and cause of death, were taken from the standardised NHS Data Dictionary (https://www.datadictionary.nhs.uk/).*

*^§^ A full list of test names is provided in Table S3. In version 2.1.0, we have added an indicator for point of care test for laboratory records.*

*^†^ Liver disease progression measurements of interest include APRI, Child-Pugh, FIB-4, METAVIR scores, fibrosis, cirrhosis, decompensated cirrhosis, and HCC.*

*^‡^ Risk factors of interest include alcohol, diabetes, HIV infection, BMI (or, Height and Weight), smoking.*

*All data classes are linked by anonymous subject identifiers.*

*^¶^ Data element ‘site provided’ for hepatitis diagnoses class, liver disease progression class, and risk factors class is used to indicate if the data were provided by the site.*

*IMDD, index of multiple deprivation decile; ICD, international classification of diseases; HBV, hepatitis B virus; HCV, hepatitis C virus; HEV, hepatitis E virus; HIV human immunodeficiency virus; BMI, body mass index; HCC, hepatocellular carcinoma; APRI, aspartate aminotransferase to platelet ratio index; FIB-4, fibrosis-4.*

Table S3. Full list of laboratory tests^§^

| **Test code** | **Test name** | **Version 2.0.0** | **Version 2.1.0** |
| --- | --- | --- | --- |
| AFPL | Alpha-fetoprotein | √ | √ |
| ALB | Albumin | √ | √ |
| ALT | Alanine Aminotransferase | √ | √ |
| AST | Aspartate Aminotrasferase | √ | √ |
| BCRE | Serum Creatinine | √ | √ |
| BTP | Total Protein | √ | √ |
| BURE | Serum Urea | √ | √ |
| CCRE | Creatinine clearance | √ | √ |
| CD4 | CD4 cell count | √ | √ |
| CFER | Ferritin | √ | √ |
| CHOL | Cholesterol | √ | Retired |
| CIRI | Insulin | √ | √ |
| CRP | C reactive protein | √ | √ |
| EGFR | eGFR | √ | √ |
| EOS | Eosinophil | √ | √ |
| GHB | Haemoglobin HBA1C | √ | √ |
| GLUC | Glucose | √ | √ |
| GSBI | Bilirubin | √ | Retired |
| HB | Haemoglobin | √ | √ |
| HBCT | Anti-Hepatitis B core | √ | √ |
| HBEB | Anti-HBe | √ | √ |
| HBEG | Hepatitis B e antigen | √ | √ |
| HBSG | Hepatitis B surface antigen | √ | √ |
| HBVD | HBV DNA | √ | √ |
| HCVA | Hepatitis C antibody | √ | √ |
| HCVL | HCV Viral Load | √ | √ |
| HCVR | HCV RNA | √ | √ |
| HDVG | HDV IgG | √ | √ |
| HDVL | HDV Viral Load | √ | √ |
| HDVM | HDV IgM | √ | √ |
| HEVM | HEV IgM | √ | √ |
| HEVG | HEV IgG | √ | √ |
| HEVP | HEV PCR | √ | √ |
| HINR | INR | √ | √ |
| HIVA | Anti-HIV | √ | √ |
| HPBG | Hepatitis B Genotype | √ | √ |
| HPCG | Hepatitis C Genotype | √ | √ |
| HPVG | Hepatitis D Genotype | √ | √ |
| HV1R | HIV 1 RNA | √ | √ |
| IGM | Immunoglobulin M | √ | √ |
| MC | Mean cell haemoglobin | √ | √ |
| MCV | Mean Cell Volume | √ | √ |
| NEUB | Neutrophil Count | √ | √ |
| NHDL | Non-HDL Cholesterol | √ | √ |
| OVITD | Vitamin D | √ | √ |
| PALK | Alkaline phosphatase | √ | √ |
| PL | Platelet count | √ | √ |
| WB | White cell count | √ | √ |
| XBIL | Bilirubin (total) | √ | √ |
| XGGT | Gamma-glutamyl transferase | √ | √ |
| ZCB | conjugated bilirubin |  | √ |
| ZCHO | Total Cholesterol | √ | √ |
| ZHDL | HDL Cholesterol | √ | √ |
| ZHDLR | Total: HDL Chol Ratio | √ | √ |
| ZTRI | Triglycerides | √ | √ |

^§^ *In Version 2.1.0, we have retired CHOL lab test code, noting that both the ZCHO and CHOL codes are used to record plasma cholesterol (i.e., total cholesterol) at different periods, and CHOL is not used anymore. We have also retired GSBI lab test code, noting that GSBI was originally used for gall stone bilirubin, which is not used anymore. We have added ZCB lab test code for conjugated bilirubin, which is of interest for viral hepatitis research.*

*HIV infection was defined by anti-HIV or HIV 1 RNA positive. HCV infection was defined by one positive of HCV viral load or HCV RNA or HCV genotype. HDV infection was defined by positive HDV antibody or detectable HDV viral load. For HEV, past infection was defined by only anti-HEV IgG positive, and acute infection was defined by anti-HEV IgM and anti-HEV IgG positive, and/or HEV PCR positive.*

Table S4. Demographics, follow up duration, and coinfection characteristics of adults with chronic HBV who died or were lost to follow-up vs. who are active in the cohort

| **Parameter** | **Active**  **n = 3864** | **Died**  **n = 261** | **Lost to follow-up**  **n=1071** | **P-value**  **died vs active** | **P-value lost to follow-up vs active** |
| --- | --- | --- | --- | --- | --- |
| Follow-up duration, years | 5.4 [3.5, 8.0] | 3.4 [1.7, 6.1] | 2.8 [1.5, 5.0] | <0.0001 | <0.0001 |
| Gender, male | 2071 (53.6) | 203 (77.8) | 567 (52.9) | <0.0001 | 0.7291 |
| Age, years | 40.0 [32.0, 50.0] | 59.0 [49.0, 69.0] | 35.0 [30.0, 45.0] | <0.0001 | <0.0001 |
| Age group, years |  |  |  |  |  |
| 18-24 | 164 (4.2) | 1 (0.4) | 82 (7.7) | <0.0001 | <0.0001 |
| 25-34 | 1063 (27.5) | 9 (3.4) | 399 (37.3) |  |  |
| 35-44 | 1165 (30.2) | 32 (12.3) | 321 (30.0) |  |  |
| 45-54 | 813 (21.0) | 52 (19.9) | 165 (15.4) |  |  |
| 55-64 | 447 (11.6) | 68 (26.1) | 60 (5.6) |  |  |
| 65-74 | 168 (4.3) | 65 (24.9) | 35 (3.3) |  |  |
| >=75 | 44 (1.1) | 34 (13.0) | 9 (0.8) |  |  |
| Ethnic groups |  |  |  |  |  |
| Asian | 1004 (26.0) | 57 (21.8) | 178 (16.6) | 0.0032 | <0.0001 |
| Black | 750 (19.4) | 55 (21.1) | 184 (17.2) |  |  |
| Mixed | 102 (2.6) | 3 (1.1) | 36 (3.4) |  |  |
| White | 929 (24.0) | 89 (34.1) | 242 (22.6) |  |  |
| Other | 386 (10.0) | 23 (8.8) | 105 (9.8) |  |  |
| Not stated | 693 (17.9) | 34 (13.0) | 326 (30.4) |  |  |
| HCV coinfection | 141 (3.6) | 42 (16.1) | 26 (2.4) | <0.0001 | 0.0628 |
| HDV coinfection | 46 (1.2) | 3 (1.1) | 7 (0.7) | 1 | 0.18 |
| HEV coinfection | 33 (0.9) | 5 (1.9) | 2 (0.2) | 0.1607 | 0.036 |
| HIV coinfection | 68 (1.8) | 3 (1.1) | 13 (1.2) | 0.6256 | 0.2676 |

Table S5. Demographic and follow-up characteristics stratified by treatment status

| Demographics | Treated  (n=828) | Untreated  (n=3485) | P-value |
| --- | --- | --- | --- |
| Follow-up duration,  years, median [IQR] | 5.0 [3.3, 7.4] | 5.2 [2.6, 8.0] | 0.309 |
| Gender = Male, n (%) | 511 (61.7) | 1899 (52.9) | <0.001 |
| Age, years, median [IQR] | 45 [35, 55] | 39 [31, 48] | <0.001 |
| Age groups, years, n (%) |  |  |  |
| 18-24 | 37 (4.5) | 195 (5.4) | <0.001 |
| 25-34 | 162 (19.6) | 1112 (30.9) |  |
| 35-44 | 207 (25.0) | 1073 (29.9) |  |
| 45-54 | 203 (24.5) | 668 (18.6) |  |
| 55-64 | 142 (17.1) | 333 (9.3) |  |
| 65-74 | 54 (6.5) | 163 (4.5) |  |
| ≥75 | 23 (2.8) | 49 (1.4) |  |
| Ethnic groups, n (%) |  |  |  |
| Asian | 241 (29.1) | 697 (19.4) | <0.001 |
| Black | 166 (20.0) | 712 (19.8) |  |
| Mixed | 26 (3.1) | 104 (2.9) |  |
| White | 206 (24.9) | 849 (23.6) |  |
| Other | 93 (11.2) | 393 (10.9) |  |
| Not stated | 96 (11.6) | 838 (23.3) |  |

IQR, interquartile range.

Table S6. Ongoing and planned studies (but not limited) using NIHR HIC HBV dataset

| Studies | Goals |
| --- | --- |
| HBV treatment eligibility and coverage | To assess HBV treatment eligibility and coverage of patients in this cohort, as well as to investigate which NICE treatment criteria and patient characteristics are associated with odds of receiving or not receiving treatment |
| Impact of COVID-19 pandemic on HBV routine surveillance | To investigate the impact of the COVID-19 pandemic on routine surveillance for adults with CHB in the UK |
| Early prediction of HBsAg loss | To apply advanced machine learning techniques to predict HBsAg loss and determine key factors associated with this endpoint |
|  | To investigate factors affecting the incidence of HCC in patients with viral hepatitis |
| HCC identification | To develop a natural language processing (NLP) pipeline to automatically identify HCC from imaging reports |
| Metabolic factors on CHB outcomes | To explore the association between metabolic risk factors and CHB outcomes |

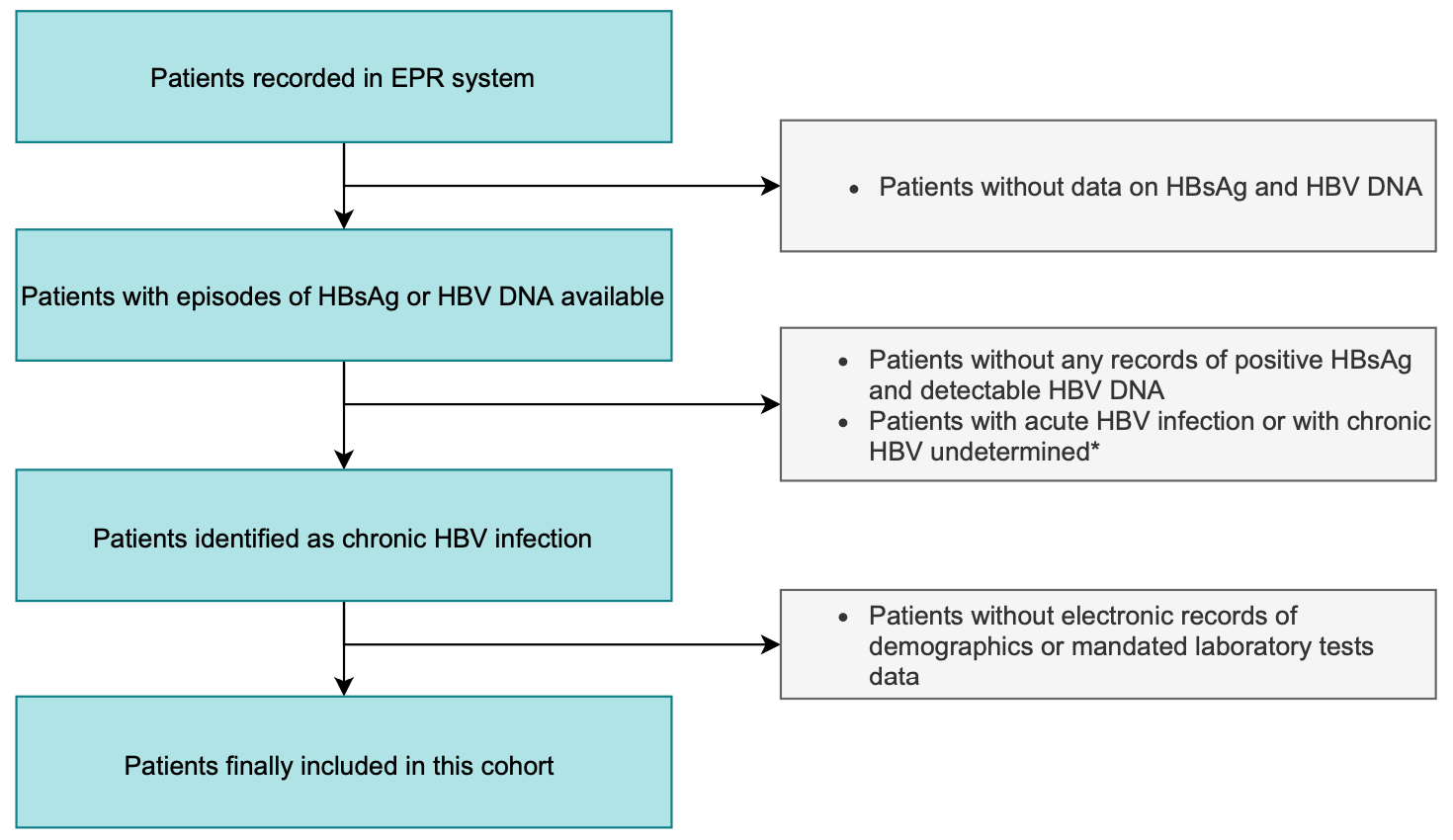

**Figure S1. Conceptual flowchart of data selection and reasons for exclusion.**

*This real process is performed by sites separately within their local data warehouses.*

**Patients with HBV infection are identified according to criteria presented in Table S1, with a change in definition implemented in June 2021.*

*EPR, electronic patient record; HBsAg, hepatitis B virus surface antigen; HBV, hepatitis B virus; DNA, deoxyribonucleic acid.*

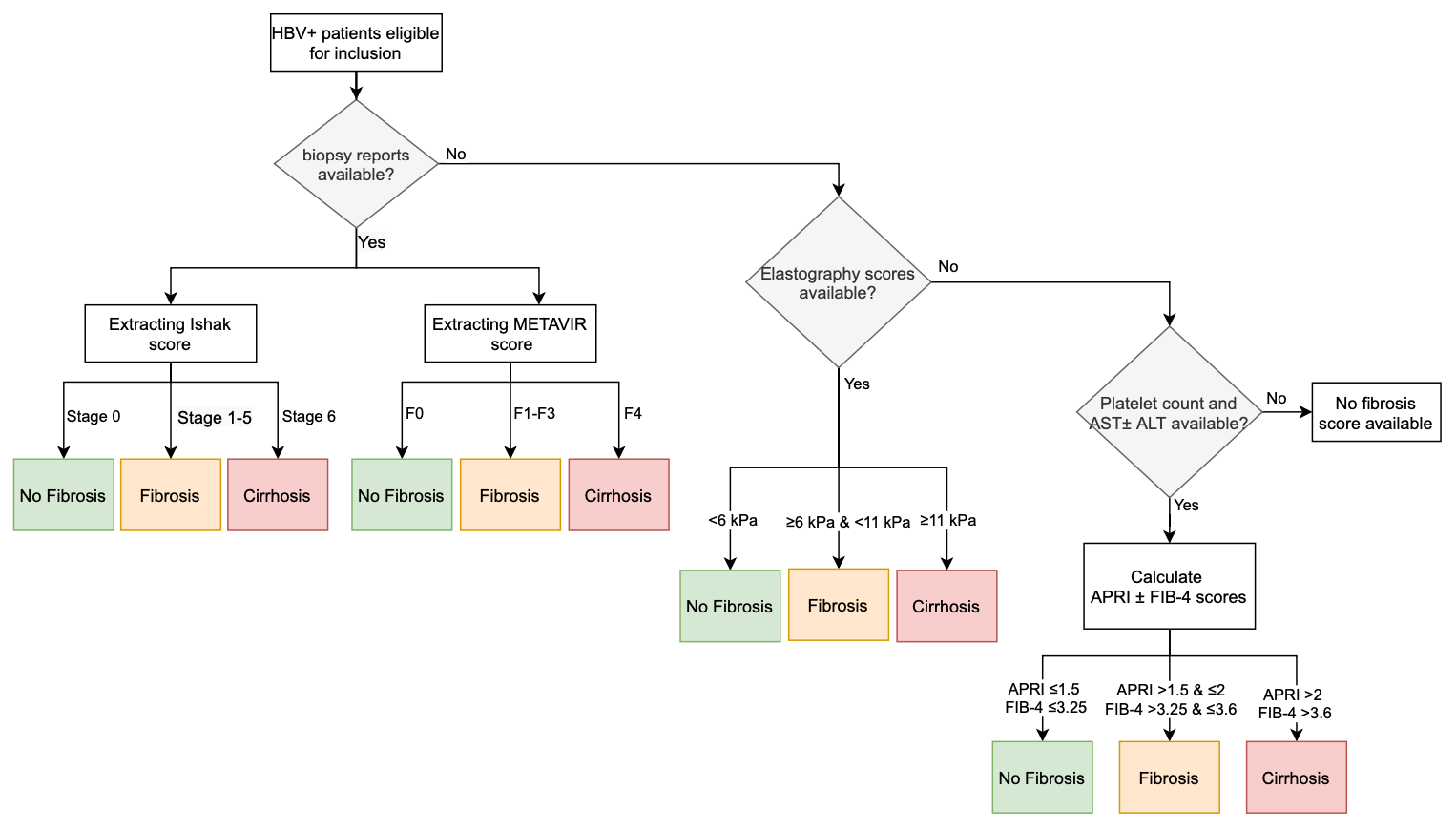

**Figure S2. The conceptual process of sequentially inferring fibrosis and cirrhosis information from various types of data.** *The current algorithm determines liver fibrosis/cirrhosis by sequentially using biopsy, elastography, or APRI/FIB-4 scores, for some patients who have two or more types of data available, the correlation of these data would need further investigated in the future studies. Also, imaging reports (ultrasound, CT, MRI) are not currently used in this algorithm due to no score scales being available in these reports, and further bioinformatics research is necessary to identify disease information.*

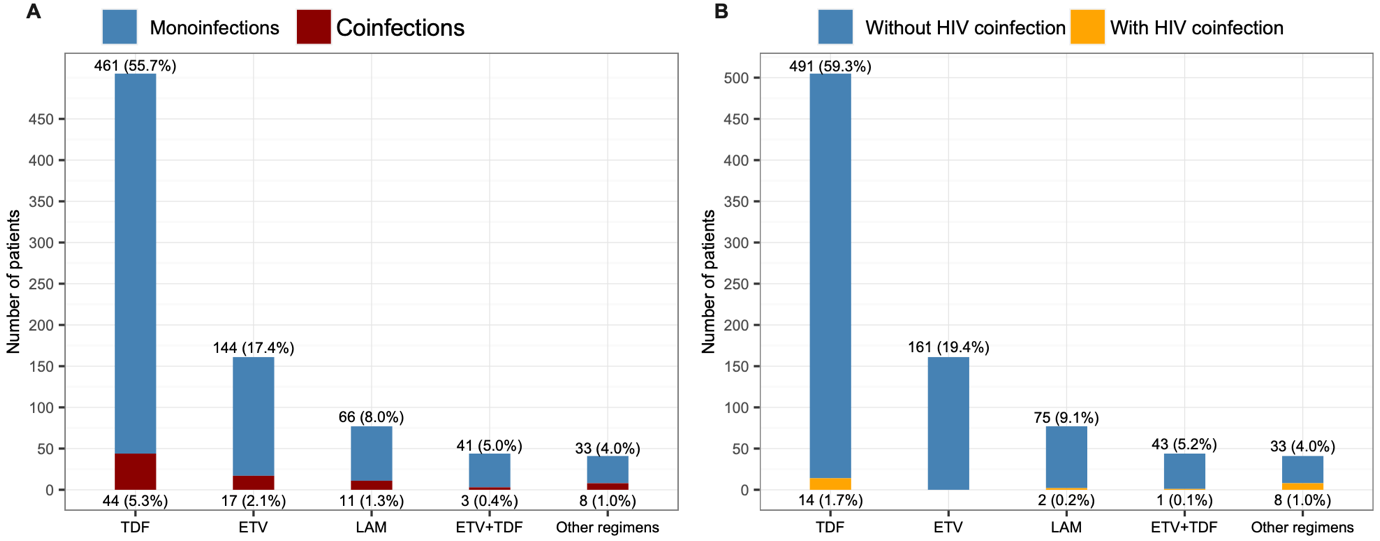

**Figure S3. Treatment regimens received by patients who were on treatment during follow-up stratified by (A) monoinfection vs. coinfections with HIV, HCV, HDV, or HEV; (B) without HIV coinfection vs. Coinfection with HIV.** *TDF, tenofovir disoproxil fumarate; ETV, entecavir; LAM, Lamivudine; ADV, adefovir dipivoxil. Treatment data are currently available from two sites. Those regimens infrequently prescribed for patients are shown together in the final column at each panel, such as combinations of LAM+TDF, ETV+LAM, ADV+LAM, or ETV+LAM+TDF. There was no episode of interferon drug recorded for the cohort although this drug is included in the data model. Each percent value was calculated as the number of patients receiving a type of regimen in each subgroup divided by the total number of patients who were on treatment.*
